# Increased galanin-galanin receptor 1 signaling, inflammation, and insulin resistance are associated with affective symptoms and chronic fatigue syndrome due to Long COVID

**DOI:** 10.1101/2024.04.25.24306334

**Authors:** Wasim Talib Mahdi Al Masoodi, Sami Waheed Radhi, Habiba Khdair Abdalsada, Menqi Niu, Hussein Kadhem Al-Hakeim, Michael Maes

**Author notes:** **Corresponding author:** Prof. Dr. Michael Maes, M.D., Ph.D. Sichuan Provincial Center for Mental Health Sichuan Provincial People’s Hospital School of Medicine University of Electronic Science and Technology of China Chengdu 610072 China. https://scholar.google.co.th/citations?user=1wzMZ7UAAAAJ&hl=th&oi=ao Highly cited author: 2003-2023 (ISI, Clarivate) ScholarGPS: Worldwide #1 in molecular neuroscience; #1/4 in pathophysiology Expert worldwide medical expertise ranking, Expertscape (December 2022), worldwide: #1 in CFS, #1 in oxidative stress, #1 in encephalomyelitis, #1 in nitrosative stress, #1 in nitrosation, #1 in tryptophan, #1 in aromatic amino acids, #1 in stress (physiological), #1 in neuroimmune; #2 in bacterial translocation; #3 in inflammation, #4-5: in depression, fatigue and psychiatry.

## Abstract

**Background:** Long COVID (LC) patients frequently suffer from neuropsychiatric symptoms, including depression, anxiety, and chronic fatigue syndrome (CFS), relabeled as the physio-affective phenome of LC. Activated immune-inflammatory pathways and insulin resistance key play a role in these physio-affective symptoms due to LC.

**Aims:** To examine the associations between the Hamilton Depression (HAMD), Hamilton Anxiety (HAMA) and Fibro-Fatigue (FF) Rating Scale scores and serum C-reactive protein (CRP), prostaglandin E2 (PGE2), galanin-galanin receptor 1 (GAL-GALR1) signaling, insulin resistance, insulin-like growth factor (IGF-1), plasminogen activator inhibitor-1 (PAI1), and damage biomarkers such as S100B and neuron-specific enolase (NSE) in 90 subjects 3-6 months after acute SARS-CoV-2 infection.

**Results:** LC patients show higher HAMD, HAMA, and FF scores, CRP, PGE2, GAL-GALR1 signaling, insulin resistance, PAI1, NSE, and S100B than participants without LC. The HAMD/HAMA/FF scores were significantly correlated with PGE, CRP, GAL, GALR1, insulin resistance, and PAI1 levels, and a composite score based on peak body temperature (PBT) – oxygen saturation (SpO2) (PBT/SpO2 index) during the acute infectious phase. A large part of the variance in the affective and CFS symptoms (33.6%-42.0%) was explained by a combination of biomarkers; the top-3 most important biomarkers were GAL-GALR1 signaling, PGE2, and CRP. Inclusion of the PBT/SpO2 index increased the prediction considerably (55.3%-67.1%). The PBT/SpO2 index predicted the increases in GAL-GALR1 signaling.

**Conclusions:** These findings suggest that the affective symptoms and CFS of Long COVID are largely the consequence of activated immune-inflammatory pathways, metabolic aberrations, and the severity of the inflammation during acute SARS-CoV-2 infection.

## Introduction

In addition to respiratory symptoms, patients who recover from coronavirus disease-2019 (COVID-19) frequently suffer from a variety of persistent neuropsychiatric disorders, such as anxiety, depression, fibromyalgia, and chronic fatigue syndrome (CFS) ^1-5^. CFS, affective symptoms (including anxiety and depression), cognitive impairments, and dyspnea are the most commonly reported symptoms in individuals with Long COVID ^6-10^. The severity of this Long COVID symptom complex resulting from long-term COVID-19 infection was significantly correlated with the peak body temperature (PBT) and oxygen saturation of peripheral blood (SpO2) (using a composite PBT/SpO2 index) during the acute inflammatory phase ^11^.

The affective symptoms and CFS associated with Long COVID are linked to oxidative stress, activated immune-inflammatory pathways, insulin resistance, SARS-CoV-2 persistence, and reactivation of specific viruses, including human herpesvirus type 6 (HHV-6) ^12, 13^. Consequently, it is valuable to investigate the potential correlation between CFS and affective symptoms resulting from Long COVID and additional metabolic or immune-inflammatory biomarkers, such as prostaglandin E2 (PGE2), insulin-like growth factor 1 (IGF-1), galanin (GAL) and its receptor type 1 (GALR1), plasminogen activator inhibitor (PAI)1, and neuronal damage markers, such as S100 calcium-binding protein B (S100B) and neuron-specific enolase (NSE).

The pulmonary tissues of individuals diagnosed with respiratory distress syndrome (ARDS) exhibit increased levels of insulin-like growth factor 1 (IGF-1), which is also referred to as somatomedin C, and the IGF-1 receptor (IGF-1R), according to research on COVID-19 infection ^14^. A hypothesis posits that inhibiting IGF-1R could potentially reduce the risk of mortality and mitigate lung impairment in patients with COVID-19-associated ARDS ^14^. IGF-1 is additionally correlated with long-term COVID-19 symptoms, including hypoxia and depression ^15, 16^.

GAL is a neuropeptide that is essential for immune-inflammatory responses ^17-20^. It is encoded by the GAL gene ^21^. Additionally, it has been observed that GAL plays a protective function within the central nervous system (CNS) ^22, 23^. Furthermore, GAL is implicated in neurocognitive disorders, depression, anxiety, pain threshold, and pain behaviors ^24-26^. The GAL receptor type 1 (GALR1) may mediate the depressive, and the GALR2 ‘antidepressant’ effects of GAL, whilst non-selective GAL receptor agonists have anxiolytic and antidepressant effects ^27, 28^.

Elevated concentrations of PGE2 have been observed in the presence of SARS-CoV-2, potentially impeding the development of immunity and obstructing the initial antiviral reaction; thus, this may promote the progression of severe diseases and recurrence of infections ^29^. By inhibiting the innate and adaptive immune systems, PGE2 facilitates the development of infections ^30, 31^. Acute inflammation is induced by PGE2 via the activation of mast cells, the differentiation of T helper 1 (Th1) cells, the proliferation of Th17 cells, and the secretion of IL-22 by Th22 cells ^32^. Long-lasting COVID symptoms, such as respiratory symptoms and chronic pain, may be influenced by PGE2 ^33^. Depression has been linked to an increase in PGE2 production ^34^.

Microscopic thrombi contribute to COVID-19 infection in addition to chronic inflammation, and oxidative stress ^35-38^. Therefore, thrombosis monitoring is a critical concern for patients with acute COVID-19 infection and maybe Long COVID. By inhibiting the activity of tissue plasminogen activator (tPA) and urokinase-like plasminogen activator (uPA), plasminogen activator inhibitor (PAI)1 is a crucial regulator of fibrinolysis ^39^. An elevation in PAI1 levels is associated with a reduction in fibrinolysis, which subsequently heightens the susceptibility to thrombosis ^40^. There are some reports that PAI is elevated in major depression ^41^.

Neuronal damage markers include S100 calcium-binding protein B (S100B) and neuronal-specific enolase (NSE) ^42, 43^. Serum S100B is a marker of clinical severity in SARS-CoV-2 infection and is involved in COVID-19 ^44^. In addition, in acute COVID-19 populations, S100B concentration correlated significantly with organ injury indicators and inflammation markers, including C-reactive protein (CRP) ^44^. In patients with acute COVID-19 infection, elevated serum NSE concentrations are correlated with disease severity ^43^ In patients with MDD and end-stage renal disease, various neuronal damage biomarkers are associated with increased levels of depression, anxiety, and CFS symptoms ^45, 46^. However, the precise mechanisms underlying the connections between neuropsychiatric symptoms due to Long COVID and biomarkers of inflammation such as PGE2, GAL-GALR1 signaling, PAI1, IGF-1, and neuronal biomarkers remain unknown. Therefore, the current investigation was conducted to explore the correlations between depression, anxiety and CFS due to Long COVID, and CRP, PGE2, GAL-GALR1 signaling, insulin resistance, PAI1, IGF1, and neuronal damage markers. Furthermore, we investigate the impact of acute inflammation induced by COVID-19 infection on the affective and CFS symptoms and biomarkers of Long COVID. The latter are measured by using a composite score based on the PBT/SpO2 index.

## 2. Subjects and Methods

### Participants

This case-control study explored differences between Long COVID patients (n=60) versus 30 controls who had previously been identified with an acute COVID-19 infection but who did not develop Long COVID. The participants were enrolled in our study during the first three months of 2023. The patients were identified based on the WHO post-COVID (long COVID) case definitions ^47^, which include the following: (a) individuals with a history of proven SARS-CoV-2 infection; (b) symptoms that persisted past the acute stage of illness or manifested during recovery from acute COVID-19 infection; (c) symptoms that lasted at least 2 months and are present 3–4 months after the onset of COVID-19; (d) patients who have no less than two symptoms that make it challenging to do everyday undertakings, for example, fatigue, headaches, trouble speaking, chest pain, a persistent cough, loss of taste or smell, emotional symptoms, mental hindrance, or fever ^47^. None of these criteria were met by the thirty controls. All individuals diagnosed with acute COVID-19 received treatment at a designated quarantine hospital located in Kerbala city, Iraq. These hospitals include Imam Al-Hussein Medical City of Kerbala, Imam Al-Hassan Al-Mujtaba Teaching Hospital, Karbala Teaching Hospital for Children, Al-Kafeel Super Specialty Hospital, and Al-Hindiyah General Hospital. Based on the presence of typical symptoms like fever, breathing issues, coughing, and loss of smell and taste, positive reverse transcription real-time polymerase chain reaction findings, and positive IgM directed to SARS-CoV-2, senior physicians and virologists diagnosed SARS-CoV-2 infection and acute COVID-19. The rRT-PCR results of all participants following the acute phase were negative.

Furthermore, the study excluded individuals who had previously suffered from diabetes mellitus or other systemic autoimmune diseases, multiple sclerosis, stroke, neurodegenerative or neuroinflammatory disorders, psoriasis, rheumatoid arthritis, inflammatory bowel disease, chronic fatigue syndrome, scleroderma, liver disease, or any other medical conditions. Additionally, the study did not include pregnant or breastfeeding women. Participants who had suffered from axis-1 disorders were excluded from participating in major depression, dysthymia, bipolar disorder, anxiety disorder, substance use disorders, and autism.

Before participating in the study, all control and patient participants, or their respective parents or legal guardians, provided written consent after receiving comprehensive information. The study received approval from the Kerbala Health Directorate-Training and Human Development Center (Document No.0030/2023) and the institutional ethics committee of the University of Kufa (1657/2023). The study adhered to both Iraqi and international ethical and privacy laws, including the International Conference on Harmonization of Good Clinical Practice, the Belmont Report, the CIOMS Guidelines, and the World Medical Association’s Declaration of Helsinki. Additionally, our institutional review board follows the International Guidelines for Human Research Safety (ICH-GCP).

### Clinical measurements

At least six months after the acute infectious phase of COVID-19, a senior pulmonologist assessed and collected sociodemographic and clinical data from Long COVID patients and their controls via semi-structured interviews. We noted which vaccines each participant had received from Sinopharm, Pfizer, or AstraZeneca. The body mass index (BMI) was calculated by dividing the kilogram of body weight by the square of the height in meters. The HAMD was used to assess the severity of depression ^48^. The same day, a senior psychiatrist assessed sociodemographic and clinical data using a semi-structured interview, and the Fibromyalgia and Chronic Fatigue Syndrome Rating (FibroFatigue or FF) scale to measure the severity of chronic fatigue syndrome (CFS)-like symptoms ^49^. The severity of anxiety was measured using the Hamilton Anxiety Rating Scale (HAM-A) ^50^. During the acute phase of infection, an experienced paramedical professional had accurately measured SpO2 levels using a state-of-the-art electronic oximeter from Shenzhen Jumper Medical Equipment Co. Ltd. Body temperature was assessed using a digital oral thermometer placed under the tongue until the beep. In this study, we collected body temperature and SpO2 levels from the patient records and used the lowest SpO2 and peak body temperature (PBT) data recorded during the acute phase of illness. Based on these assessments, we have calculated a new index taking into account the decrease in SpO2 and the increase in body temperature, represented as the z transformation of PBT (z PBT) - z SpO2 ^11^.

### Assays

Five milliliters of fasting blood samples were taken at around 9:00 a.m. Following a ten-minute waiting period, the clotted blood samples were centrifuged at 1200Xg for five minutes. Later, the serum was divided and placed into three Eppendorf tubes. Hemolyzed samples were not included in the study. These tubes were then frozen at -80 °C until thawed for assays. We used an ELISA technique to quantify serum human GAL, GALR1, PAI1, IGF-1, insulin, PGE2, NSE, and S100B using ready-for-use ELISA kits supplied by Nanjing Pars Biochem Co., Ltd. (Nanjing, China). To reflect GALR1 signaling we computed a z unit-based composite score as z GAL + z GALR1 (labeled as GAL+GALR1). The intra-assay coefficient of variation (CV) of all ELISA kits was <10.0%. We used sample dilutions for samples containing analytes with high concentrations. CRP levels in human serum were assessed using CRP latex slide assays (Spinreact^®^, Barcelona, Spain). Fasting serum glucose levels were determined spectrophotometrically using a ready-to-use kit provided by Spinreact^®^ (Barcelona, Spain).

The Homeostasis Model Assessment 2 (HOMA2) calculator^©^ (Diabetes Trials Unit, University of Oxford; https://www.dtu.ox.ac.uk/homacalculator/download.php) was used to calculate insulin sensitivity (HOMA2%S), and insulin resistance (HOMA2IR) from the fasting serum insulin and glucose levels.

### Statistical analysis

The distribution types of the results group were investigated using the Kolmogorov-Smirnov test. The mean ± standard deviation was used to express the results for normally distributed variables. Analysis of variance (ANOVA) was used to examine differences in continuous variables between groups and contingency tables (χ^2^-test) to examine associations between nominal variables. The Mann-Whitney U (MWU) test was used to compare biomarkers between the two study groups. Pearson’s product-moment correlation coefficients were used to investigate the links between variables. The relationships between categories and biomarkers were examined using multivariate general linear model (GLM) analysis (followed by tests of between-subject effects) that took into consideration confounding variables, such as sex, age, and BMI. Using clinical and biomarker levels as explanatory variables in manual and automatic stepwise (p-to-enter of 0.05 and p-to-remove of 0.06), multiple regression analysis was used to delineate the significant biomarkers predicting the neuropsychiatric scores. We examined the effects of all biomarkers, with or without the z PBT – z SpO2 ratio and examined the associations also in the restricted study group of Long COVID patients. We calculated the standardized beta coefficients for each significant explanatory variable using *t* statistics with the exact value of p, as well as the model F statistics and total variance explained (R^2^) as effect size. The tests were two-tailed, and statistical significance was defined as a value of p of 0.05. We used IBM SPSS 28 for Windows to examine the data.

## Results

### Sociodemographic and clinical data

**Table 1** shows that there were no significant differences among groups in age, sex ratio, marital status, residency, employment, education, BMI, TUD, vaccination type, and period from the acute infection to inclusion in the study between the two study groups. The duration of the acute infection was greater in Long-COVID than in the control group. The Long COVID group showed a significantly higher PBT than the control group. During the acute phase of COVID infection, SpO2 was lower in the Long COVID group compared with the controls. The Long COVID patients displayed significantly greater scores on the FF, HAMA, and HAMD scores than controls.

**Table 1.** Sociodemographic and clinical variables in subjects with and without Long COVID. All results are shown as mean (±SD) or as ratios; the former are analyzed using analysis of variance, ratios are analyzed using analysis of contingency tables (χ2 test). MWU: Mann-Whitney U test. BMI: body mass index, PBT: peak body temperature, HAMA: Hamilton Anxiety Rating Scale, HAMD: Hamilton Depression Rating Scale, FF: Fibro-Fatigue scale, SpO2: oxygen saturation, Vaccination A, PF, S: vaccination with AstraZeneca, Pfizer, or Sinopharm.

| Variables | No Long COVID<br>(n=30) | Long COVID<br>(n=60) | F/$\chi^2$ | df | p |
| --- | --- | --- | --- | --- | --- |
| Age (years) | 35.8±7.5 | 37.0±9.4 | 0.38 | 1/88 | 0.538 |
| Sex (Female/Male) | 10/20 | 27/33 | 1.12 | 1 | 0.289 |
| Body mass index (kg/m<sup>2</sup>) | 28.30±4.06 | 28.49±5.06 | 0.03 | 1/88 | 0.858 |
| Marital status (No/Yes) | 5/25 | 7/53 | 0.43 | 1 | 0.511 |
| Rural / Urban | 6/24 | 13/47 | 0.03 | 1 | 0.855 |
| Employment (No/Yes) | 5/25 | 13/47 | 0.31 | 1 | 0.576 |
| Education (years) | 12.9±4.9 | 11.5±5.0 | 1.68 | 1/88 | 0.198 |
| Tobacco use disorder (No/Yes) | 18/12 | 37/23 | 0.23 | 1 | 0.878 |
| Duration of the acute infectious phase (days) | 8.6±5.7 | 15.3±9.1 | 13.44 | 1/88 | <0.001 |
| Time from acute infection to study inclusion (months) | 16.0±5.9 | 17.3±6.1 | 0.81 | 1/88 | 0.370 |
| SpO<sub>2</sub> (%) | 92.97±3.48 | 80.33±12.87 | 27.74 | 1/88 | <0.001 |
| PBT (°C) | 38.45±0.70 | 39.10±0.92 | 11.39 | 1/88 | 0.001 |
| Vaccination (Non/A/PF/S) | 9/2/14/5 | 24/4/28/4 | 2.54 | 3 | 0.467 |
| Total FF score | 8.0±3.5 | 41.1±11.2 | MWU | - | <0.001 |
| Total HAMA score | 4.9±1.8 | 25.1±9.3 | MWU | - | <0.001 |
| Total HAMD total | 5.1±2.8 | 20.98±7.1 | MWU | - | <0.001 |

### Biomarkers in Long COVID

**Table 2** shows the results of multivariate GLM analysis which examined the association between the biomarkers and diagnosis (Long COVID versus controls) while adjusting for age, sex, and BMI. Tests for between-subject effects showed that there were significant associations between the diagnosis and all biomarkers, except IGF-I. **Table 3** shows the mean biomarker values in Long COVID patients versus non-Long COVID controls. Glucose, insulin, HOMA2IR, CRP, PAI1, PGE2, NSE, GAL, GALR1, and S100B were significantly increased in Long COVID patients in comparison with the controls. There was a significant decrease in HOMA2%S in Long COVID compared with controls, whereas IGF-1 was not significantly different between both groups.

**Table 2.** Results of multivariate GLM analysis that examines the association among the biomarkers and the diagnosis of Long COVID versus no Long COVID, after covarying for age, body mass index (BMI), and sex. * Processed in log transformation, ** processed in square root transformation, CRP: C-reactive protein, S100B: calcium-binding protein B, PAI1: Plasminogen activator inhibitor-1, HOMA2IR: Homeostasis Model Assessment (HOMA) 2 insulin resistance index, HOMA2%S: HOMA insulin sensitivity percentage, NSE: neuron-specific enolase, IGF-1: Insulin-Like Growth Factor-I, PGE2: prostaglandin E2, GALR1: Galanine receptor 1.

| Tests | Biomarkers | Covariates | F | df | p |
| --- | --- | --- | --- | --- | --- |
| **Multivariate** | All biomarkers | Diagnosis | 6.20 | 12/74 | <0.001 |
|  |  | Sex | 0.83 | 12/74 | 0.621 |
|  |  | Age | 1.924 | 12/74 | 0.045 |
|  |  | BMI | 2.13 | 12/74 | 0.025 |
| **Between-subject effects** | Glucose | Diagnosis | 14.72 | 1 | <0.001 |
|  | Insulin | Diagnosis | 21.99 | 1 | <0.001 |
|  | HOMA2%S | Diagnosis | 15.75 | 1 | <0.001 |
|  | HOMA2IR | Diagnosis | 24.64 | 1 | <0.001 |
|  | CRP* | Diagnosis | 9.83 | 1 | 0.002 |
|  | PAI1 * | Diagnosis | 8.05 | 1 | 0.006 |
|  | IGF-1 * | Diagnosis | 2.54 | 1 | 0.115 |
|  | PGE2 | Diagnosis | 16.38 | 1 | <0.001 |
|  | NSE* | Diagnosis | 4.76 | 1 | 0.032 |
|  | Galanin | Diagnosis | 9.59 | 1 | 0.003 |
|  | GALR1 * | Diagnosis | 8.41 | 1 | 0.005 |
|  | S100B** | Diagnosis | 5.09 | 1 | 0.027 |

**Table 3:** Estimated marginal mean (SE) values of the biomarker data in subjects with and without Long COVID. The p-values are obtained by the GLM analysis shown in Table 2. Transformations were used where needed, namely *logarithmic; ** square root. For abbreviations: see Table 2.

| **Variables** | **No Long COVID**<br>**(n=30)** | **Long COVID**<br>**(n=60)** | **p** |
| --- | --- | --- | --- |
| Glucose (mM) | 5.14±0.32 | 6.00±1.18 | <0.001 |
| Insulin (pM) | 72.70±24.72 | 106.84±34.70 | <0.001 |
| HOMA2%S | 86.96±44.58 | 56.35±26.48 | <0.001 |
| HOMA2IR | 1.36±0.47 | 2.05±0.66 | <0.001 |
| CRP* (mg/l) | 3.50±1.35 | 6.51±4.81 | 0.002 |
| PAI1* (ng/ml) | 2.79±0.68 | 4.44±2.96 | 0.006 |
| IGF-1* (ng/ml) | 20.70±5.00 | 25.39±18.17 | 0.115 |
| PGE2* (pg/ml) | 17.20±4.66 | 38.27±29.56 | <0.001 |
| NSE* (ng/ml) | 14.89±7.26 | 17.98±8.10 | 0.032 |
| Galanin* (pg/ml) | 48.2±15.3 | 87.7±69.0 | 0.003 |
| GALR1* (pg/ml) | 54.9±11.0 | 89.7±74.5 | 0.005 |
| S100B** (pg/ml) | 199.8±164.2 | 431.7±521.4 | 0.027 |

### Intercorrelation matrix among neuropsychiatric symptoms and biomarkers

The intercorrelation matrix among the FF, HAMA, and HAMD scores, the biomarkers and z PBT – z SpO2 ratio are presented in **Table 4**. The clinical scores were significantly correlated with glucose, insulin, HOMA2IR, GAL, and GALR1. The HAMA and HAMD scores were significantly correlated with CRP, PAI1, PGE2, and S100B, whilst there were significant and inverse associations with the HOMA2%S. The FF score was significantly and positively correlated with HOMA2%S, CRP, PAI1, IGF-1, and S100B (all positively correlated), and inversely with HOMA2%S. The composite z PBT – z SpO2 score is significantly correlated with the neuropsychiatric scores and IGF-1.

**Table 4.**
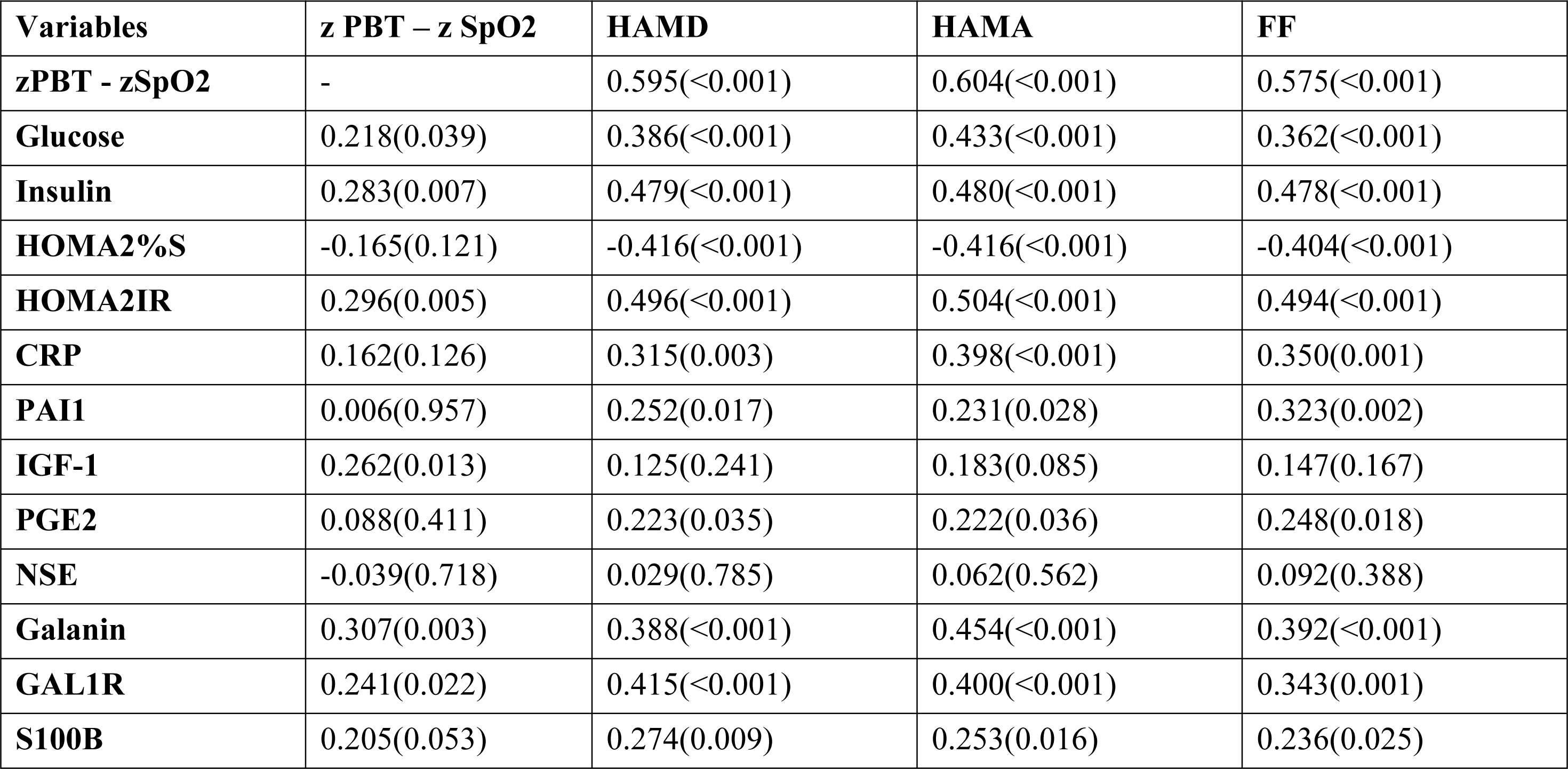
Correlation matrix among neuropsychiatric test scores and an index of increased peak body temperature and lowered peripheral blood oxygen saturation (zPBT - zSpO2) during acute COVID-19 infection, and biomarkers of Long COVID. Z: z-score, CRP: C-reactive protein, S100B: calcium-binding protein B, HOMA2IR: Homeostasis Model Assessment (HOMA) 2 insulin resistance index, HOMA2%S: HOMA insulin sensitivity percentage, NSE: neuron-specific enolase, IGF-1: Insulin-Like Growth Factor-I, PGE2: prostaglandin E2, PAI1: Plasminogen Activator Inhibitor Type-1, GALR1: Galanine receptor 1, HAMA: Hamilton Anxiety Rating Scale, HAMD: Hamilton Depression Rating Scale, FF: fibro fatigue scale.

### Results of the multiple regression analysis

**Table 5** presents the results of multiple regression analyses with the neuropsychiatric domains as dependent variables and biomarkers as explanatory variables. Regression #1.1 shows that 33.6% of the variance in the FF symptom score was explained by the regression on CRP, PGE2, and GAL+GALR1. Regression #1.2 shows that 42.0% of the variance in the HAMA score was explained by the regression on CRP, PGE2, and the composite GAL+GALR1 score. Regression #1.3 shows that 38.4% of the variance in the HAMD score was explained by the regression on CRP, PGE2, female sex, and GAL+GALR1. **Figure 1** shows the partial regression of the HAMA score on the GAL+GALR1 composite score. The findings of multiple regression analyses using biomarkers and the z SpO2 -z PBT ratio as explanatory factors are shown in the same Table. Regression #2.1 shows that 55.3% of the variance in the FF score was explained by the regression on CRP, PAI1, HOMA2IR, z PBT – z SpO2, and GAL+GALR1. **Figure 2** shows that partial regression plot with the total scores of FF as the dependent variable and the severity of inflammation during the acute phase as an explanatory variable. Regression #2.2 shows that 67.1% of the variance in the HAMA score is explained by the regression on glucose, GAL, CRP, and GALR1 (all positively), and sex and SpO2 (both negatively). Regression #2.3 shows that 57.1% of the variance in the HAMD was explained by the regression on z PBT – z SpO2, HOMA2IR, GALR1, CRP, and sex.

**Figure 1.**
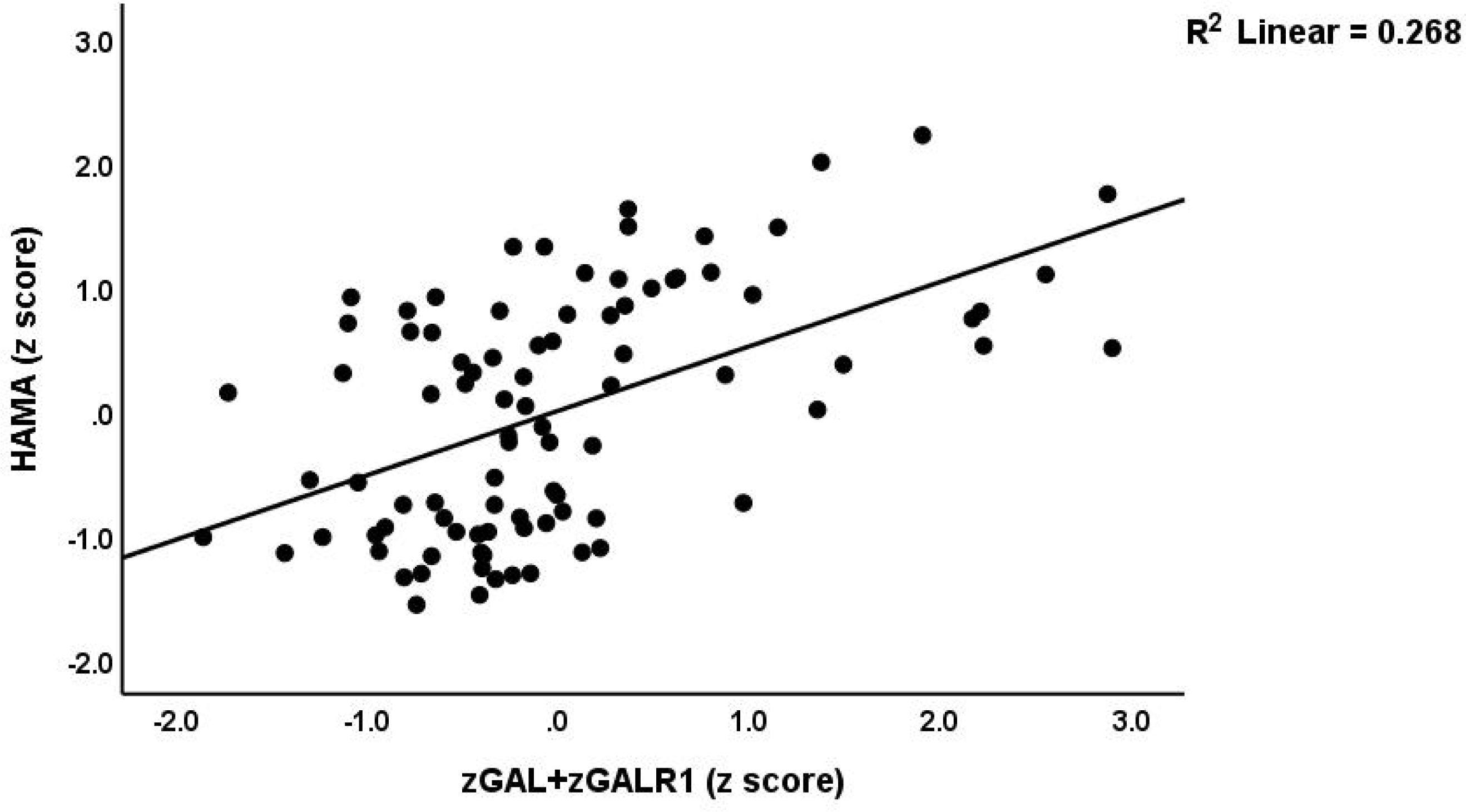
Partial regression plot with the scores of Hamilton Anxiety Rating Scale (HAMA) score as dependent variable on an integrated index (conceptualized as a z unit-based composite) of galanin (GAL) + GAL receptor 1 (GALR1) signaling (denoted as zGAL+zGALR1), after adjusting for age, sex, and body mass index; p<0.001.

**Figure 2.**
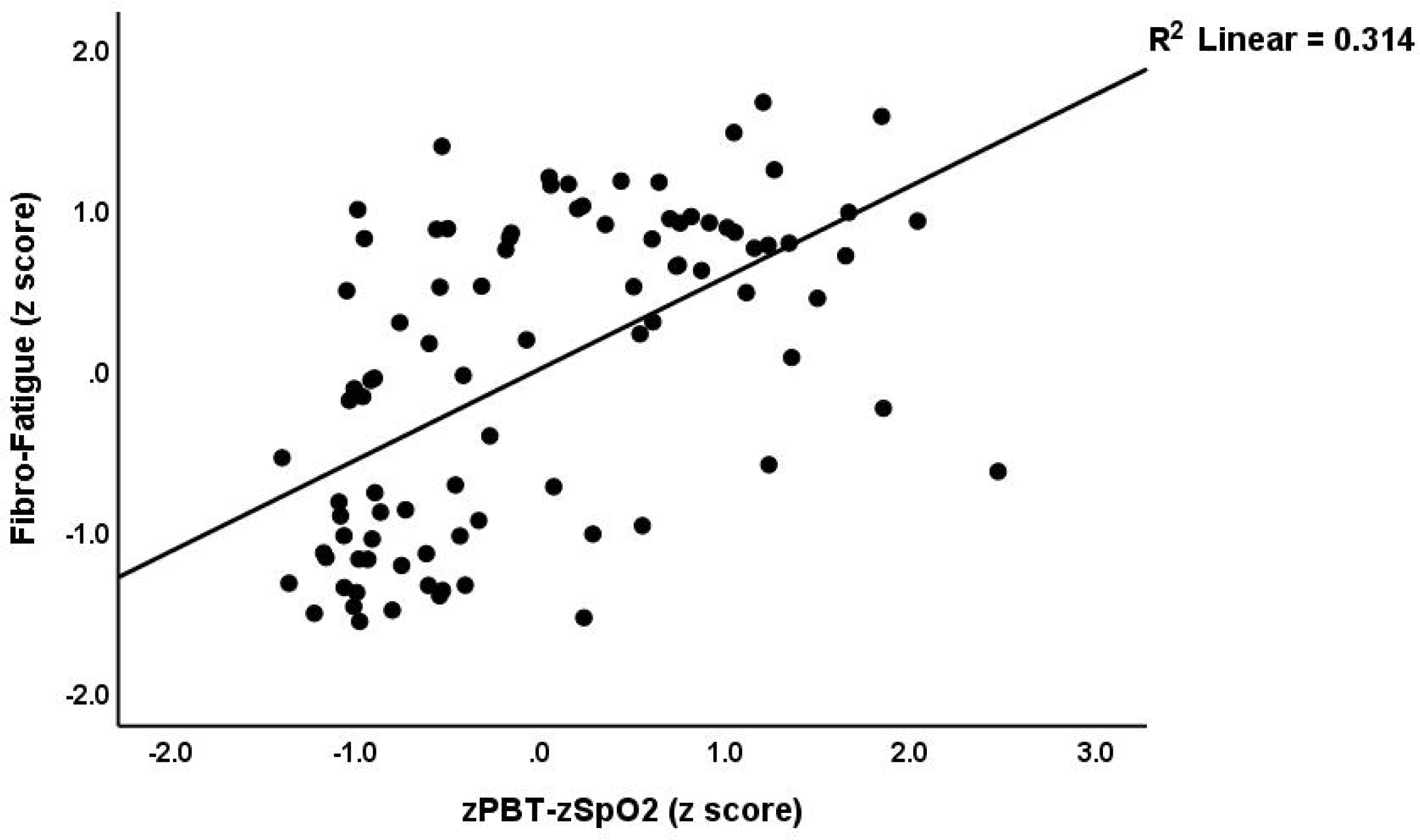
Partial regression plot with the Fibro-Fatigue scale (FF) score as dependent variable and an integrated index of peak body temperature (PBT) and oxygen saturation (SpO2) (zPBT-zSpO2) as explanatory variable (after adjusting for the effects of age, sex and body mass index); p<0.001.

**Table 5.** Results of multiple regression analysis with neuropsychiatric rating scale scores as dependent variables and biomarkers as explanatory variables. HAMA: Hamilton Anxiety Rating Scale, HAMD: Hamilton Depression Rating Scale, FF: Fibro-Fatigue scale, CRP: C-reactive protein, PGE2: prostaglandin E2, GALR1: Galanin receptor 1, GAL+GALR1: z unit composite score of galanin and GALR1, PBT – SpO2: z composite score based on peak body temperature – oxygen saturation (SpO2), indicating the severity of inflammation during the acute phase. HOMA2IR: Homeostasis Model Assessment (HOMA) 2 insulin resistance index, HOMA2%S: HOMA insulin sensitivity percentage.

| Regression | Explanatory variables | $\beta$ | t | p | F<sub>model</sub> | df | p | R<sup>2</sup> |
| --- | --- | --- | --- | --- | --- | --- | --- | --- |
| #1.1. FF | **Model** |  |  |  | 14.52 | 3/86 | <0.001 | 0.336 |
|  | GAL+GALR1 | 0.426 | 4.78 | <0.001 |  |  |  |  |
|  | CRP | 0.249 | 2.76 | 0.007 |  |  |  |  |
|  | PGE2 | 0.221 | 2.48 | 0.015 |  |  |  |  |
| #1.2. HAMA | **Model** |  |  |  | 20.72 | 3/86 | <0.001 | 0.420 |
|  | GAL+GALR1 | 0.492 | 5.91 | <0.001 |  |  |  |  |
|  | CRP | 0.292 | 3.46 | 0.001 |  |  |  |  |
|  | PGE2 | 0.190 | 2.28 | 0.025 |  |  |  |  |
| #1.3. HAMD | **Model** |  |  |  | 13.27 | 4/85 | <0.001 | 0.384 |
|  | GAL+GALR1 | 0.492 | 5.68 | <0.001 |  |  |  |  |
|  | CRP | 0.198 | 2.26 | 0.027 |  |  |  |  |
|  | PGE2 | 0.205 | 2.38 | 0.020 |  |  |  |  |
|  | Sex | -0.192 | -2.25 | 0.027 |  |  |  |  |
| #2.1. FF | **Model** |  |  |  | 20.81 | 5/84 | <0.001 | 0.553 |
|  | PBT-SpO2 | 0.419 | 5.20 | <0.001 |  |  |  |  |
|  | HOMA2IR | 0.235 | 2.89 | 0.005 |  |  |  |  |
|  | PAI1 | 0.180 | 2.29 | 0.025 |  |  |  |  |
|  | CRP | 0.175 | 2.31 | 0.024 |  |  |  |  |
|  | GAL+GALR1 | 0.169 | 2.06 | 0.042 |  |  |  |  |
| #2.2. HAMA | **Model** |  |  |  | 28.23 | 6/83 | <0.001 | 0.671 |
|  | SpO2 | -0.441 | -6.48 | <0.001 |  |  |  |  |
|  | Glucose | 0.270 | 4.16 | <0.001 |  |  |  |  |
|  | GAL | 0.221 | 3.25 | 0.002 |  |  |  |  |
|  | CRP | 0.219 | 3.37 | 0.001 |  |  |  |  |
|  | GALR1 | 0.194 | 2.91 | 0.005 |  |  |  |  |
|  | Sex | -0.155 | -2.42 | 0.018 |  |  |  |  |
| #2.3. HAMD | **Model** |  |  |  | 22.33 | 5/84 | <0.001 | 0.571 |
|  | PBT - SpO2 | 0.430 | 5.61 | <0.001 |  |  |  |  |
|  | HOMA2IR | 0.280 | 3.66 | <0.001 |  |  |  |  |
|  | GALR1 | 0.241 | 3.21 | 0.002 |  |  |  |  |
|  | CRP | 0.162 | 2.21 | 0.030 |  |  |  |  |
|  | Sex | -0.156 | -2.17 | 0.033 |  |  |  |  |
| #3.1. FF | **Model** |  |  |  | 9.40 | 1/58 | 0.003 | 0.139 |
|  | PBT – SpO2 | 0.373 | 3.07 | 0.003 |  |  |  |  |
| #3.2. HAMA | **Model** |  |  |  | 9.93 | 4/55 | <0.001 | 0.419 |
|  | PBT – SpO2 | 0.360 | 3.42 | 0.001 |  |  |  |  |
|  | GAL+GALR1 | 0.348 | 3.31 | 0.002 |  |  |  |  |
|  | CRP | 0.244 | 2.37 | 0.021 |  |  |  |  |
|  | Glucose | 0.213 | 2.07 | 0.043 |  |  |  |  |
| #3.3. HAMD | **Model** |  |  |  | 8.96 | 3/56 | <0.001 | 0.324 |
|  | PBT – SpO2 | 0.301 | 2.63 | 0.011 |  |  |  |  |
|  | GAL+GALR1 | 0.270 | 2.39 | 0.020 |  |  |  |  |
|  | HOMA2%S | -0.266 | -2.35 | 0.022 |  |  |  |  |

The same Table shows the results of multiple regression analyses performed in the restricted study group of patients with Long COVID. Regression #3.1 shows that a considerable part of the variance in FF-total (13.9%) was explained by the regression on z PBT – z SpO2. A significant part of the variance of the HAMA total score (41.9%) can be explained by the regression on z PBT-z SpO2, GAL+GALR1, and CRP (regression #3.2). Regression #3.3 shows that 32.4% of the variance in the HAMD score was explained by the regression on z PBT – z SpO2 and GAL+GALR1 (positively associated) and HOMA%S (inversely associated). **Figure 3** shows the partial regression plot with the scores of HAMD as the dependent variable and the composite GAL+GALR1 as explanatory variable.

**Figure 3.**
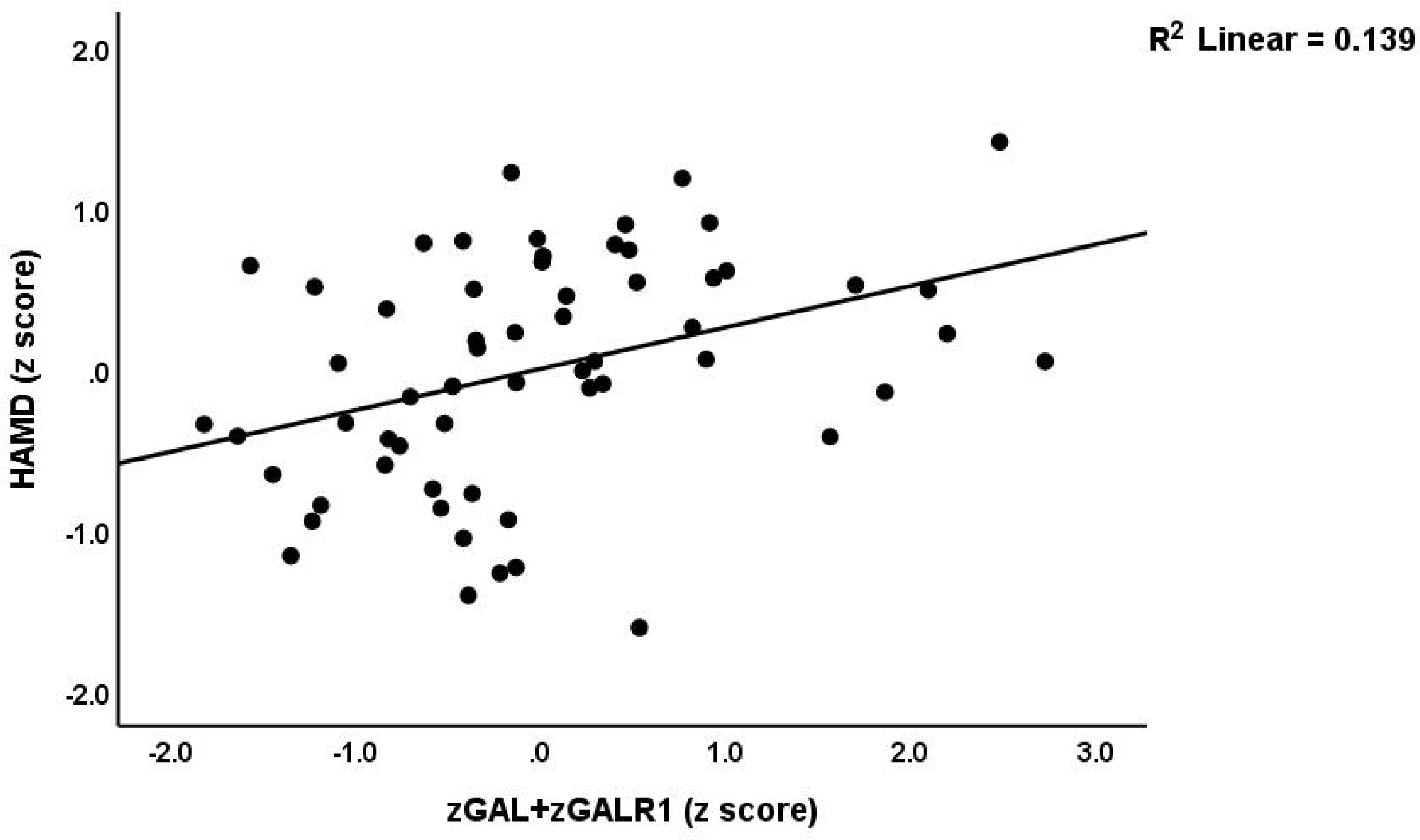
Partial regression plot with the scores of Hamilton Depression Rating Scale (HAMD) as dependent variable and the z unit-based composite z GAL+GALR1 (i.e. galanin and Galanin receptor 1) as explanatory variable (after adjusting for age, sex, and body mass index); p<0.01. This regression was performed in the restricted study sample of patients with Long COVID.

## Discussion

### The physio-affective phenome of Long COVID

One significant discovery from this study is that Long COVID is marked by higher scores on affective and CFS rating scales. Our previous research has demonstrated that both the acute infectious phase and Long COVID are associated with heightened affective and CFS symptoms ^11, 51, 52^. Given the simultaneous expression of these elevations in both affective and physiosomatic symptom domains, we have reclassified this symptom profile as the physio-affective phenome of acute COVID-19 infection and Long COVID ^11, 52, 53^. In addition, the current study found that the various aspects of the physio-affective phenome of Long COVID were strongly influenced by decreased oxygen saturation (SpO2) and increased peak body temperature (PBT) that were recorded during the initial infectious stage. These findings align with previous studies that have shown a strong correlation between the severity of inflammation during the acute phase of SARS-CoV-2 infection and the physio-affective phenome of Long COVID ^11^. These findings indicate that the Long COVID physio-affective phenome is influenced by the immune-inflammatory processes that occur during the acute infection ^11, 12, 53, 54^.

### Biomarkers of Long COVID

The present study reveals that Long COVID exhibits a range of biomarkers that indicate immune-inflammatory responses (such as increased PGE2 and CRP), metabolic changes (including increased glucose, insulin, HOMA2IR, and PAI1), lung and/or neuronal damage (as evidenced by increased NSE and S100B), and GAL-GALR1 signaling (with increased GAL and GALR1 levels).

As mentioned in the Introduction, recent findings suggest that Long COVID is characterized by heightened immune-inflammatory pathways and oxidative stress. This is supported by the presence of increased CRP, the NLRP3 inflammasome, elevated lipid peroxidation, and reduced antioxidant defenses ^12, 55-57^. The current research indicates that individuals with Long COVID exhibit elevated levels of serum PGE2 and CRP compared to the control group, providing additional evidence of the involvement of immune-inflammatory mechanisms. In previous studies, it was found that during the early phase of SARS-CoV-2 infection, there was an increase in serum PGE2 levels. These elevated levels were shown to hinder the body’s initial antiviral defense mechanisms and the development of immunity, which in turn led to more severe cases of COVID-19 infection and an increased risk of multiple infections ^29^. Therefore, it is plausible that elevated PGE2 levels during the initial stages of infection could contribute to the development of Long COVID by heightening susceptibility to the persistence and reactivation of the SARS-CoV-2 virus ^13^. In addition, the persistence of PGE2 from the acute phase to the Long COVID phase could potentially heighten the susceptibility to viral persistence or reactivation. PAI1 has effects on various aspects of health, including thrombosis, inflammatory responses, and cardiovascular disease ^58^. PAI1 has the potential to trigger inflammatory processes in alveolar epithelial cells ^59^. Therefore, PAI1 may potentially contribute to the inflammatory mechanisms involved in COPD ^59^. Furthermore, it is worth noting that higher levels of pro-inflammatory cytokines such as IL-6 and TNF-α may stimulate the production of PAI1 ^60^.

Elevated GAL levels could potentially contribute to the immune mechanisms involved in the pathophysiology of Long COVID. GAL has the potential to exert immune-suppressive effects, as demonstrated in studies conducted by de Medeiros et al. (2022) ^61^ and Oliveira-Volpe et al. (2020) ^62^. Nevertheless, the GAL-GALR1 signaling pathway has been found to decrease the levels of cyclic adenosine monophosphate (cAMP), as demonstrated in a study by Koller et al. (2019) ^63^. cAMP possesses anti-inflammatory properties by suppressing the production of pro-inflammatory cytokines and promoting the expression of IL-10 ^64^. According to Koller et al. (2019), the production of proinflammatory cytokines/chemokines is increased by GAL in M1 polarized macrophages. Interestingly, an opposite response is observed in unpolarized macrophages ^63^. Therefore, the activation of M1 macrophage polarization in Long COVID can potentially lead to more profound pro-inflammatory effects of GAL signaling ^62, 63^. Moreover, the GAL/GALR1 system potentially plays a role in the fundamental mechanisms of Long COVID by influencing endocrine systems ^20, 65-67^. GAL is present in sensory and cholinergic nerve fibers in the airways, indicating a potential role of this peptide in influencing airway vascular and secretory function. In addition, the GAL distribution indicates potential effects on airway, vascular, and secretory functions in the respiratory tract of mammals ^68^. It appears that GAL smooth muscle has a significant impact on the regulation of gastrointestinal and respiratory smooth muscle function ^69^. Thus, it is possible to propose a potential connection between GAL and the inflammatory processes associated with pulmonary diseases ^70^.

It has been reported that Long COVID is marked by the development of insulin resistance, as shown by an elevated HOMA2IR index ^71, 72^. COVID-19, being a respiratory tract infection, has been found to affect various cell types and can result in extrapulmonary consequences ^73, 74^. Research findings indicate that SARS-CoV-2 could invade and reproduce within pancreatic β-cells, which are responsible for producing insulin. This can lead to a disruption in insulin production and secretion, contributing to the metabolic disturbances observed in individuals with COVID-19. Several studies have provided evidence supporting this connection ^75-78^. An examination of infected pancreatic cell cultures revealed that SARS-CoV-2 effectively manipulates the ribosomal machinery within these cells, as confirmed by the transcriptome analysis conducted ^79^. Additional mechanisms involve the influence of inflammation and glucocorticoids or the formation of micro-thrombotic lesions that lead to abnormalities in blood flow within the pancreatic islets ^80, 81^. In addition, GAL has the ability to reduce the secretion of insulin and somatostatin, as demonstrated in a study conducted by Oliveira Volpe et al. (2020) ^62^.

There is a mounting body of evidence suggesting that S100B plays a significant role in inflammatory processes triggered by DAMP and AGE-RAGE stress ^44^. S100B can induce the release of pro-inflammatory cytokines, which are frequently implicated in lung inflammation. This mechanism is believed to be similar to the one potentially involved in COVID-19, as suggested by Piazza et al. (2013)^82^ and Mete et al., (2021) ^83^. Levels of NSE exhibit an increase during the acute phase of COVID-19, potentially serving as an additional indicator of disease progression ^84, 85^. The NSE protein is primarily found in the cytoplasm of neurons and neuroendocrine cells ^84^. NSE has a shown potential in aiding the diagnosis, treatment, and monitoring of various lung injuries, both acute and chronic ^86, 87^. It has also been studied in relation to solitary pulmonary nodules ^88^ and infectious lung diseases like tuberculosis ^89^. Therefore, the heightened NSE levels observed in Long COVID could potentially suggest the presence of lung damage and/or damage to the peripheral or central nervous system.

### Biomarkers and the physio-affective phenome

One notable discovery from this study is that a sizable portion of the variation in the physio-affective phenome was accurately predicted by a combination of biomarkers, including GAL-GALR1 signaling, CRP, PGE2, and z PBT - z SpO2. These biomarkers emerged as the top predictors in the analysis. Consequently, the physio-affective symptoms of Long COVID are largely influenced by immune-inflammatory pathways, such as inflammatory mediators (CRP, PGE2), metabolic pathways linked to immune-inflammatory networks (insulin resistance, PAI1), biomarkers of lung or neuronal injury (NSE, S100B), and GAL-GALR1 signaling, which regulates immune, metabolic, and hormonal responses.

In a previous study, it was demonstrated that the physio-affective phenome of Long COVID can be predicted by certain indicators of immune activation, such as CRP and the NLRP-3 inflammasome, along with related pathways of oxidative stress ^12^. According to the authors, they found that approximately 60% of the variation in the physio-affective symptoms of Long COVID, as measured by HAMD, HAMA, and FF scores, could be attributed to elevated oxidative toxicity, reduced antioxidant levels, and an increased z PBT – z SpO2 ratio. The results obtained in the current study show a similar level of explained variance. Hence, a sizable portion of the variation in affective and CFS symptoms of Long COVID can be ascribed to immune-inflammatory and associated pathways.

A recent study demonstrated that elevated levels of CRP and IgM antibodies to HHV-6-duTPase, along with IgG antibodies targeting HHV-6 and IgA antibodies targeting activin-A, can be indicative of the affective symptoms associated with Long COVID ^90^. Based on the findings of the study, it was observed that CFS symptoms could be most accurately predicted by CRP, IgG to HHV-6-duTPase, IgM to activin-A, IgM to SARS-CoV-2, and IgA-activin-A. Based on inference, it can be concluded that the persistent activation of immune-inflammatory pathways, SARS-CoV-2 persistence, and viral reactivation (HHV-6) significantly influence the affective and CFS symptoms of Long COVID. Activation of immune-inflammatory pathways may result from the acute inflammatory condition of the acute infection (as determined by PBT and SpO2 values), the persistent SARS-CoV-2 infection and subsequent viral reactivation, and the elevated PGE2 levels seen in acute infections ^91^ and Long COVID (this study).

In previous studies, researchers have found a connection between PAI1 and the development of major depression and depressive-like behaviors in animal models of depression ^41^. Research suggests that the GALR1 receptor plays a role in the depressive effects of GAL, as indicated by studies conducted by Rajarao et al. (2007) ^27^, and Kuteeva et al. (2008) ^28^. Therefore, the interaction between PAI1 and the GAL-GALR1 signaling could potentially exacerbate the immune-metabolic-hormonal abnormalities in Long COVID. S100B plays a crucial role as a danger signal and serves as a key regulator of M1 macrophage inflammation ^92^. Research suggests that alpha-enolase, which has the potential to transform into NSE, plays a role in triggering inflammatory processes that could potentially harm neuronal structures ^93^. In addition, it is worth noting that NSE has been found to have proinflammatory effects and the potential to contribute to the degradation of the extracellular matrix and neuronal injuries ^93^. Therefore, elevated levels of S100B and NSE could potentially play a role in the development of lung injuries, the continuous immune-inflammatory response, and/or damage to neurons. Thus, the activated immune-inflammatory pathways in Long COVID may be further exacerbated by GAL-GALR1 signaling, PAI1, S100B, and NSE. The results of the present study provide evidence that immune-inflammatory activation is a significant factor in the physio-affective phenome of Long COVID.

Furthermore, a prior study demonstrated that the physiological and affective characteristics of Long COVID are linked to higher levels of fasting blood glucose and insulin, as well as an elevated HOMA2IR index, suggesting the development of insulin resistance ^12^. The current findings support the idea that insulin resistance contributes to the affective and CFS of Long COVID. In addition, the study conducted by Al-Hakeim et al. (2023) found a correlation between the onset of insulin resistance and the severity of inflammation during the acute phase of illness ^94^. The authors of the study found that heightened insulin resistance plays a role in worsening and contributing to the immune-related neurotoxicity that leads to the physiological and emotional symptoms of Long COVID.

## Conclusions

Patients with Long COVID exhibit elevated levels of HAMD, HAMA, and FF scores, as well as increased CRP, PGE2, GAL-GALR1 signaling, insulin resistance, PAI1, NSE, and S100B compared to individuals without LC. The HAMD/HAMA/FF scores showed significant correlations with several factors such as PGE, CRP, GAL, GALR1, insulin resistance, PAI1 levels. Additionally, a composite score based on PBT and SpO2 was found to be associated with these clinical scores. A significant portion of the variability in the affective and CFS symptoms, ranging from 33.6% to 42.0%, can be attributed to a combination of the immune and metabolic biomarkers. Among these biomarkers, the GAL-GALR1 signaling, PGE2, and CRP emerged as the top three most influential factors. The addition of the PBT/SpO2 composite score significantly improved the accuracy of the prediction, increasing it from 55.3% to 67.1%. The findings indicate that the affective and physiosomatic symptoms of Long COVID are primarily caused by the activation of immune-inflammatory pathways, metabolic abnormalities, and the intensity of inflammation during the initial SARS-CoV-2 infection. Immune biomarkers such as PGE and the NLRP-3 inflammasome, oxidative stress pathways, GAL-GAL-R1, and metabolic pathways including insulin resistance and PAI1 are new drug targets to prevent and treat Long COVID and its physio-affective symptoms.

## Ethics statement

The study received approval from the Kerbala Health Directorate-Training and Human Development Center (Document No.0030/2023) and the institutional ethics committee of the University of Kufa (1657/2023).

## Human and animal rights

The study was conducted ethically under the World Medical Association Declaration of Helsinki and in compliance with Iraqi, international, and privacy legislation. Additionally, under the Declaration of Helsinki, The Belmont Report, CIOMS Guidelines, and the International Conference on Harmonization in Good Clinical Practice (ICH-GCP), our IRB abides by the International Guidelines for the Protection of Human Research Subjects.

## Consent for publication

Before participating in this study, each subject provided written informed consent.

## Author’s contributions

All the contributing authors have participated in the preparation of the manuscript.

## Data availability statement

The database created during this investigation will be provided by the corresponding author (MM) upon a reasonable request once the authors have thoroughly used the data set.

## Acknowledgments

We acknowledge the assistance of the workers at the Imam Al-Hussein Medical City of Kerbala, Imam Al-Hassan Al-Mujtaba Teaching Hospital, Karbala Teaching Hospital for Children, Alkafeel Super Speciality Hospital, and Al-Hindiyah General Hospital in the Kerbala Governorate of Iraq for their assistance in gathering sample material. We appreciate Maytham Abdulameer Al Maamory and Ammar Abbas Neamh, senior pulmonologists in the Clinic of Respiratory Medicine, as well as the highly qualified staff of the hospital’s internal labs, for their assistance in estimating the levels of biomarkers.

